# Open-source solution for evaluation and benchmarking of large language models for public health

**DOI:** 10.1101/2025.03.20.25324040

**Authors:** Laura Espinosa, Djilani Kebaili, Sergio Consoli, Kyriaki Kalimeri, Yelena Mejova, Marcel Salathé

## Abstract

Large language models (LLMs) have demonstrated remarkable capabilities in various natural language processing tasks, including text classification, information extraction, and sentiment analysis. However, most existing benchmarks are general-purpose and lack relevance for domain-specific applications such as public health. To address this gap, the objective of this study is to develop and test an open-source solution enabling intuitive, easy and rapid benchmarking of popular LLMs in public health contexts.

An LLM prediction prototype was developed, supporting multiple popular LLMs. It enables users to upload datasets, apply prompts, and generate structured JSON outputs for LLM tasks. The application prototype built with R and the Shiny library facilitates automated LLM evaluation and benchmarking by computing key performance metrics for categorical and variables. We tested these prototypes on four public health use cases: stance detection towards vaccination in tweets and Facebook posts, detection of vaccine adverse reaction in BabyCenter forum posts, and extraction of epidemiological quantitative features from the World Health Organization Disease Outbreak News.

Results revealed high variability in LLM performance depending on the task, dataset, and model, with no single LLM consistently outperforming others across all tasks. While larger models generally excelled, smaller models performed competitively in specific scenarios, highlighting the importance of task-specific model selection.

This study contributes to the effective LLM integration in public health by providing a structured, user-friendly and scalable solution for LLM prediction, evaluation and benchmarking. Our findings underline the relevance of standardised and task-specific evaluation methods and model selection, and the use of clear and structured prompts to improve LLM performance in domain-specific use cases.

## 1. Introduction

Large language models (LLMs) are a type of language model that use neural networks with billions of parameters and are trained on a vast amount of unlabeled data using a self-supervised approach [1] based on Transformers [2]. This has revolutionised their application in sophisticated and general-purpose language tasks, where traditional natural language processing techniques and other language models struggled to achieve high performance without burdensome and costly fine-tuning approaches. LLMs excel in tasks such as text translation and summarisation, data extraction and sentiment analysis, which are relevant across various domains, including healthcare and public health [3–5]. Furthermore, previous studies have shown the increased performance of LLMs in comparison to other machine learning approaches in public health use cases such as extracting the stance towards public health interventions [6–8].

Since 2022, the number of newly released LLMs has increased exponentially [9], requiring continuous benchmarking to ensure the adoption of cutting-edge technology. However, existing benchmarking standards are not always applicable to public health because they rely on general-purpose datasets and tasks, which do not capture the specialised terminology, unstructured data formats and complexity typical of public health use cases.

Several studies have assessed the performance of specific LLMs on concrete health or public health tasks [4,6,10,11]. A recent systematic review on testing and evaluation of healthcare applications of LLMs between 2022 and 2024 concluded that current evaluations are fragmented and insufficient [9]. Additionally, Reddy (2023) proposed a framework for evaluating LLMs in healthcare, which included human assessment of results and emphasised the need to evaluate other aspects such as utility, ethics, and functionality [12].

Several frameworks have been developed to evaluate large language models across multiple tasks and domains. For example, HELM (Holistic Evaluation of Language Models) provides a standardized evaluation across scenarios such as question answering, summarization, and classification, using a range of metrics including accuracy, calibration, and fairness [13]. Evidently-AI offers tools for monitoring and evaluating machine learning models, including LLMs, with features such as dashboarding and performance tracking across versions [14]. These efforts have contributed to more transparent and consistent model evaluation in general-purpose settings. However, these frameworks are primarily designed for technical users and require programmatic setup or integration into development pipelines. They do not currently offer interfaces or benchmarks specifically tailored to public health tasks or workflows.

For general benchmarking of LLMs performance in language skills, mathematical problem-solving and conversational abilities, standardised tests have been developed to compare different models [14]. While these tests promote a harmonised approach to LLMs evaluation, these are not always available for domain-specific application and are not always directly applicable to public health. Given these challenges, a user-friendly tool for LLMs prediction, evaluation and benchmarking is essential to ensure successful implementation in the public health field. Furthermore, when developing a prototype, testing it with real examples is crucial for assessing its performance before full deployment.

The objective of this study is to develop and test an open-source solution that enables an intuitive, easy and rapid benchmarking of popular LLMs for public health use cases. Following the key tasks that LLMs are expected to perform, we selected four public health use cases to evaluate the performance of specific LLMs in their respective objectives.

## 2. Materials and methods

This study consisted of three major pillars: (i) LLMs prediction prototype for categorical and numerical variables, (ii) Shiny app prototype for LLMs evaluation and benchmarking, and (iii) four different public health use cases for testing the first two pillars.

### 2.1. LLM prediction prototype

We built an LLM prediction prototype based on Ruby 3.4.2 as the scripting language and Ruby on Rails 8.0.1 and HTML for the web application. The application has three main components: datasets, prompts and LLMs.

The datasets must be CSV files with at least one column named “##_MSG_##_TEXT” which is the text from which the LLM will predict the desired categorical or numerical variable (e.g., stance towards vaccination from social media posts or number of cases reported in a short document). The file can contain other columns which will be ignored by the prototype. The users can upload as many datasets as needed. It will also allow images as input in the next releases.

Each dataset can have one or more experiments which is the combination of a specific prompt and single LLM (Fig 1). The application loops for each row of the designated column in the dataset, adding the specified prompt (provided by the user in plain text) and sending these to the Application Programming Interface (API) endpoint of the specified LLM. The prototype forces the LLM to give a JSON file as output; however, it is recommended to include clear instructions as well in the prompt on the requirement to provide a JSON file as output and on the keys or variables that should be provided for the prediction.

**Figure 1.**
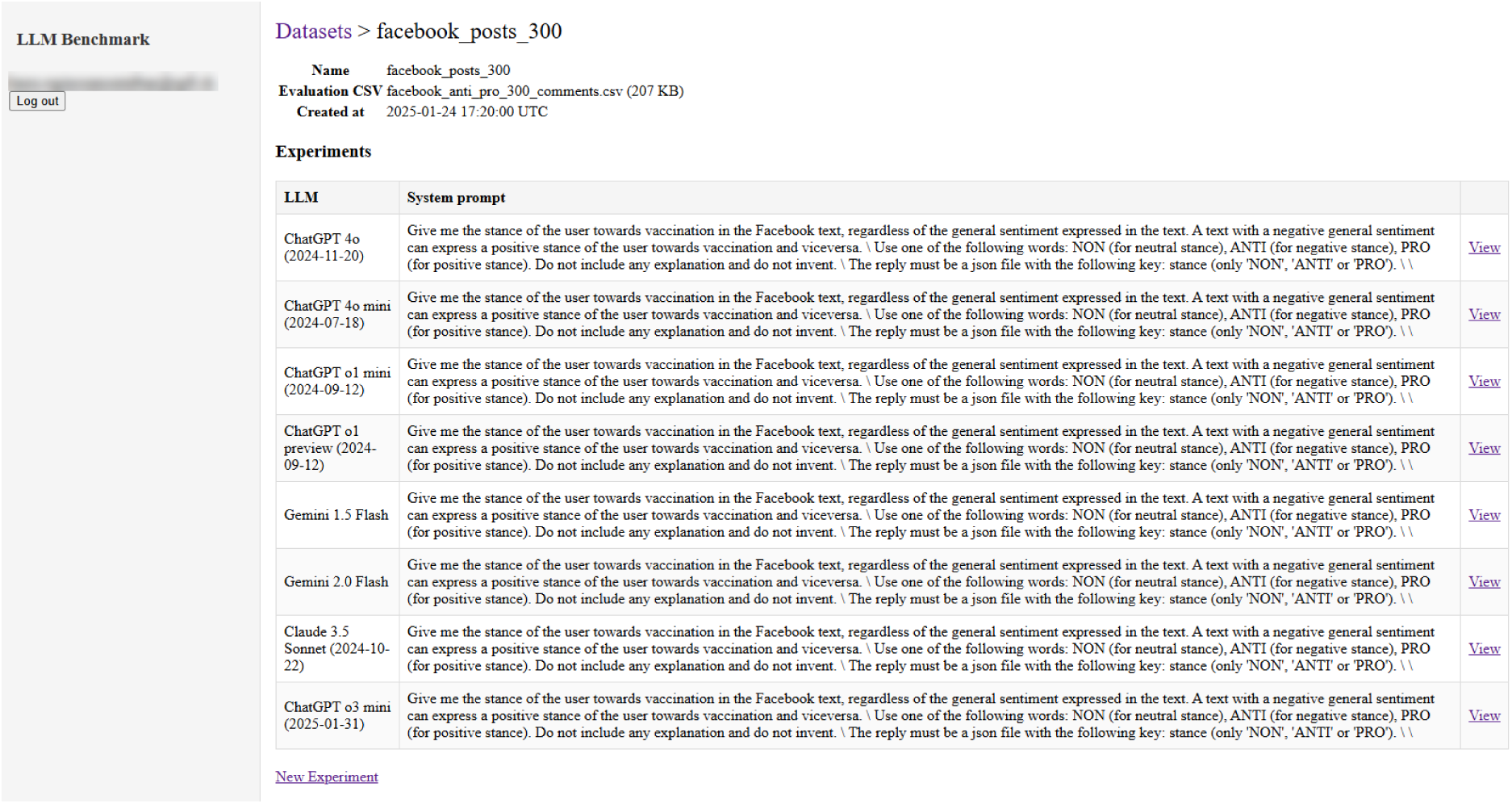
Screenshot of the LLM prediction prototype with the list of experiments for one of the public health use cases included in this study.

To date, the prototype includes the following LLMs: Generative Pre-trained Transformer (GPT) 4o (endpoint 2024-11-20), GPT 4o mini (endpoint 2024-07-18), GPT o1 preview (endpoint 2024-09-12), GPT o1 mini (endpoint 2024-09-12) and GPT o3 mini (endpoint 2025-01-31) as commercial models, and Gemini 1.5 Flash, Gemini 2.0 Flash, Deepseek R1 (Distilled Qwen 32B via Hugging Face, hereafter referred to as Deepseek R1 D), and Claude 3.5 Sonnet (endpoint 2024-10-22) as proprietary models. The specific cost associated with the use of a commercial LLM needs to be covered by each user, and the corresponding tokens for accessing the related model endpoints should be inserted in the application accordingly. Considering the changes in pricing and specific conditions depending on the account, users should refer to the latest terms and conditions of the LLMs.

### 2.2. Shiny app prototype for LLM evaluation and benchmarking

We have developed a Shiny app for LLMs evaluation and benchmarking, after predicting the value of the variable(s). This app has been built with R Studio 2024.04.2 and R version 4.5.0, and it is available as open-access at https://github.com/digitalepidemiologylab/llm_benchmarking.

The app has three pages: instructions, and LLMs evaluation and benchmarking of categorical and numerical variables. The instructions page includes information on the tool, requirements and step-by-step instructions. The other two pages contain two main panels: left panel, to include all the parameters needed to evaluate the LLMs results and the steps required to process the data and calculate the performing metrics according to the variable type; and right panel, showing the outcome and results of each step. Each page has unique parameters and results, thus information from one page is not automatically transferred to another page.

The app requires two files: a CSV file with the labelled variable and a column with a unique identifier for each entry, and a CSV or JSON file with the LLM predicted variable and the same unique identifier for each entry as the first file. The second file can be the output from the LLM prediction prototype or from any other tool.

The parameters required to be provided by the user are: paths to the dataset; LLM predictions; folder where to store the evaluation and benchmarking results; specific LLM that has been used; name of the target and LLM predicted columns; and name of the specific experiment.

The app includes a series of steps for evaluating and benchmarking LLMs. It begins by importing the labeled and the LLM prediction datasets. These datasets are then merged using a unique identifier for each entry. For categorical variables, the evaluation is done by calculating the confusion matrix which includes sensitivity, specificity, positive predictive value, negative predictive value, precision, recall, F1 score, detection rate, detection prevalence, and balanced accuracy for each class, as well as overall accuracy. Additionally, a 95% confidence interval is provided for the overall accuracy with a one-sided test to determine if the accuracy was better than selecting the majority class. For numerical variables, the evaluation includes accuracy, root-mean-square error (RMSE), and mean absolute error (MAE) as metrics.

### 2.3. Public health use cases

Four different public health use cases were selected to test the functionality and usability of the LLM prediction and Shiny app for LLM evaluation and benchmarking prototypes. These use cases were: (i) predicting stance towards vaccination in English tweets, (ii) predicting stance towards vaccination in Facebook posts, (iii) detecting vaccine adverse reactions from the digital parenting forum BabyCenter United States (US), and (iv) extracting relevant epidemiological information from the World Health Organization (WHO) Disease Outbreak News (DON). The prompts used for each public health use case are available in the supporting information (S1 Text) and were extracted from previous publications using the same data when available.

#### 2.3.1. Stance towards vaccination in English tweets

The objective of this use case is to predict the stance toward vaccination (neutral, negative or positive) in 1,000 random English-language tweets on vaccination collected between 2nd of December, 2019 and 11th of March, 2022. The stance towards vaccination may differ from the general sentiment of the tweet. The gold standard dataset was labelled by a convenience sample of four experts from the EPFL Digital Epidemiology Lab: two public health experts, one expert annotator, and one expert in administration as a representative of the general population [6].

#### 2.3.2. Stance towards vaccination in Facebook comments

The objective of this use case is to predict the stance towards vaccination in Facebook comments from pages supportive or contrary to vaccination. Facebook pages containing the keywords ‘vaccine’, ‘vaccines’ or ‘vaccination’ were queried using the Facebook Graph API between January 2012 and June 2019 by Beiró *et al*., 2023 [11]. The gold standard dataset is a random sample of 300 Facebook comments (150 from pages supportive to vaccination and 150 from pages contrary to vaccination) labelled by nine annotators familiar with the moral foundation’s theory.

#### 2.3.3. Detecting vaccine adverse reactions from BabyCenter US forum

The objective of this use case is to detect the mention of vaccine adverse reactions (yes, no) from BabyCenter US forum posts and comments. BabyCenter is a digital platform for new or expectant parents that has more than 34 million monthly users worldwide from its eight websites [15]. Posts and comments with the keyword ‘vaccine’ from 11th of March, 2008 to 26th of April, 2019 were queried in the site search function by Betti *et al*., 2021 [10] in compliance with the Terms of Use. The gold standard dataset was a random sample of 300 comments labelled by seven annotators familiar with the vaccination debate.

#### 2.3.4. Extracting relevant features from World Health Organisation (WHO) Disease Outbreak News (DONs)

The objective of this use case is to extract relevant epidemiological features (i.e., affected country, date of the event, reported number of confirmed cases and deaths, and reported disease or pathogen) from WHO DONs. WHO DONs fulfill the requirement of the Article 11.4 of the International Health Regulation (2005) to make information on acute public health events available if there is a need for disseminating authoritative and independent information [16]. It is not an exhaustive list of public health events to which WHO is responding to. We selected a random sample of 300 WHO DONs between 30th of July, 1996 and 2nd of April, 2019. The used gold standard dataset was labelled by the Information Centre for International Health Protection at Robert Koch Institut [17,18].

#### 2.3.5. Exploratory data analysis of the public health use cases

We have analysed the LLMs predictions for five categorical variables and two numerical variables from the four public health use cases. All categorical variables were imbalanced and numerical variables did not have a normal distribution (Fig 2). The majority classes were “neutral” for stance towards vaccination in tweets and Facebook posts and “no” for mentions of vaccine adverse reactions in BabyCenter forum posts. There were 89 unique affected countries reported in WHO DONs and the most frequently reported were Democratic Republic of the Congo (COD) and Guinea (GIN), which each of them were reported in 6.3% of the news, and Liberia (LBR), Nigeria (NGA) and Uganda (UGA), which each of them were reported in 4% of the news. There were 42 unique diseases reported in WHO DONs and the most frequently reported were Ebola virus and cholera, which were reported in 22.0% and 19.7% of the news, respectively. The total number of cases reported in all WHO DONs included in this study had a median of 181 and an interquartile range (IQR) of 616,011. The total number of deaths reported in all WHO DONs included in this study had a median of 39 and an IQR of 3,881.

**Figure 2.**
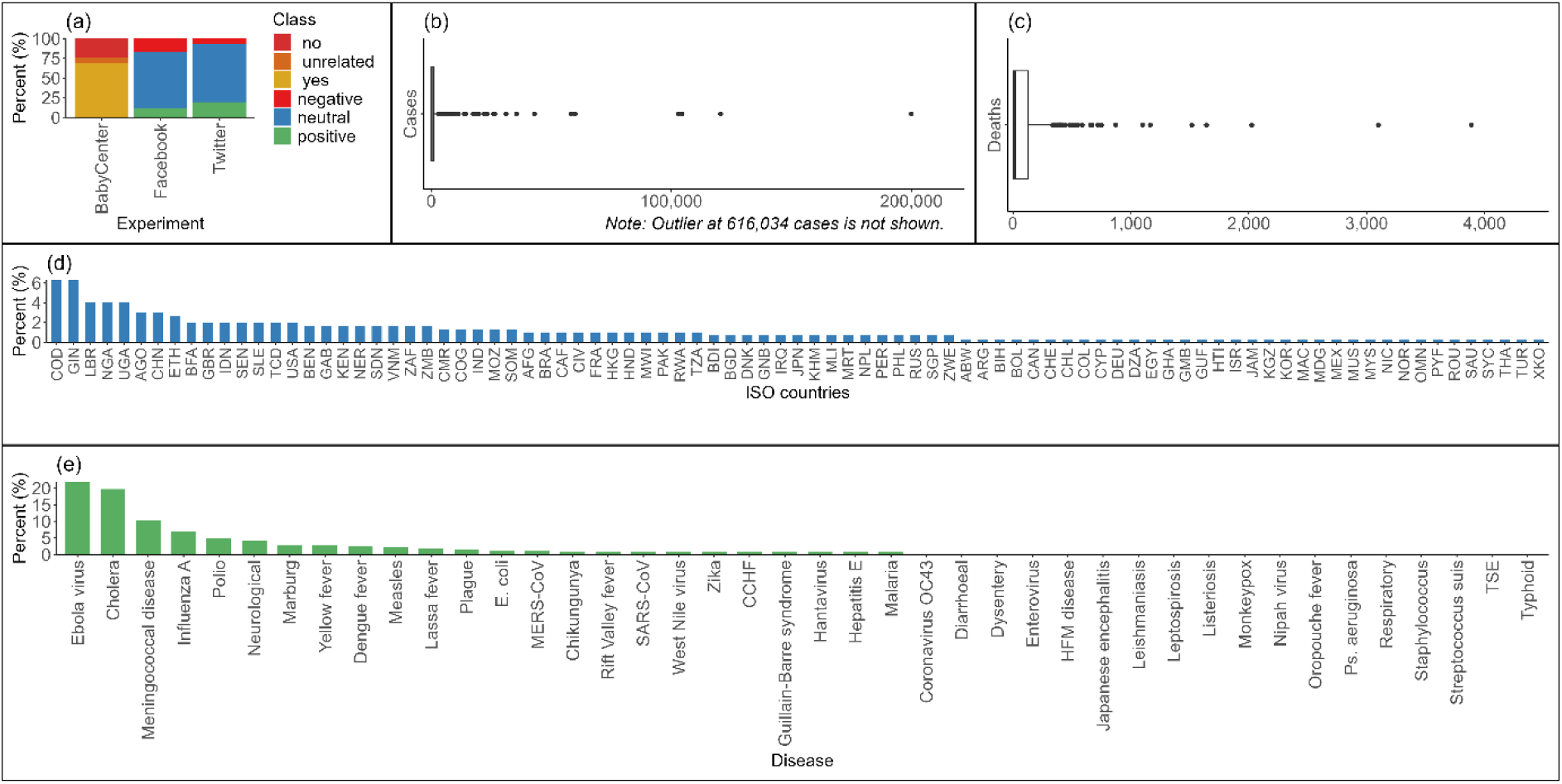
Distribution of classes and values of the seven variables analysed. (a) Class distribution for predictions from tweets, Facebook comments and BabyCenter posts, (b) Boxplot of cases reported in WHO DONs, (c) Boxplot of deaths reported in WHO DONs, (d) Affected countries reported in WHO DONs, and (e) Diseases or pathogens reported in WHO DONs.

### 2.4. Preprocessing of LLM results

We performed two data cleaning processes on the LLM results: one to extract predictions from the reasoning provided by the LLM where applicable, and another to manually revise the LLM results for WHO DONs due to complexity and lack of established ontologies for the disease. We used R 4.5.0 and text mining for the former and manual selection for the latter.

We used the Shiny app prototype to assess the LLM results compared to the annotated datasets for each use case.

### 2.5. Code and data sharing

Data and code used for the LLMs prediction prototype and evaluation and benchmarking can be found in the LLM benchmarking repository (https://github.com/digitalepidemiologylab/llm_benchmarking). In addition, data can be found at https://doi.org/10.6084/m9.figshare.29860595.v2.

## 3. Results

### 3.1. Design considerations for Usability of the LLM prediction prototype and Shiny app

The two prototypes (LLM prediction and Shiny app for LLM evaluation and benchmarking) are open-source tools that are designed with usability and accessibility in mind to allow their use by public health experts. The repositories of the prototypes include information on its use and all necessary code to execute them.

Although no usability testing or metrics were collected, several design principles were incorporated to facilitate the adoption by users, including public health professionals with limited technical expertise. The LLM prediction prototype is designed to provide outputs in a harmonised JSON format and displays the prompts used for each experiment, supporting reproducibility and reusability across different LLMs or datasets. Predictions can be easily downloaded after the experiment is finalised. The Shiny app prototype was designed to accommodate different types of inputs, following a predefined structure, including, but not only limited to, the JSON files from the LLMs prediction prototype. It includes modifiable parameters to support different experiments. Its user interface is implemented in R Shiny, requiring minimal setup and technical requirements. The application runs locally and does not rely on external APIs or authentication, reducing complexity for new users.

### 3.2. Preprocessing of LLM results

All outputs for all use cases from the model Deepseek R1 D required data cleaning to extract the predictions and delete the reasoning provided by the model.

The nomenclature used by LLMs for the diseases and/or pathogen was not harmonised among diseases/pathogens and LLMs in the WHO DONs. The LLM predictions were manually corrected by a public health expert and co-author of this study to align the nomenclature of diseases and/or virus with the labelled dataset (e.g., Ebola, Ebola virus disease or Ebola haemorrhagic fever) and to select the extracted features from the initial or main country affected. For example, the WHO DON on Ebola virus disease outbreak in West Africa [19] was first detected in Guinea but soon after, cases were reported in Sierra Leone and Liberia as well.

Between 41 and 60% of the WHO DONs required this manual revision and, from these, between 92.7 and 100% referred to the nomenclature used for the diseases and the remaining revisions referred to selecting the epidemiological information for the main affected country. The diseases which required manual alignment with the annotated dataset in all LLMs were: cholera, dengue fever, Ebola virus, enterovirus, hepatitis E, human coronavirus OC43, influenza A, Marburg fever, meningococcal disease, Oropouche fever, polio, SARS-CoV and Zika virus disease.

### 3.3. LLM evaluation and benchmarking

#### 3.3.1. Overview of accuracy across public health use cases

The overall accuracy of the LLM predictions differed depending on the LLM, target variables and experiment (Fig 3). In predicting vaccine adverse reactions in BabyCenter posts, the accuracy was very stable among LLMs with results showing statistically significant improvements over just selecting the majority class. In predicting stance towards vaccination in Facebook posts and tweets, the accuracy varied according to the used LLM with higher variability for tweets and worse accuracy in most of the predictions than just selecting the majority class. The best performing LLMs were Gemini 2.0 and GPT-4o for Facebook posts and GPT o1 mini and GPT o3 mini for tweets.

**Figure 3.**
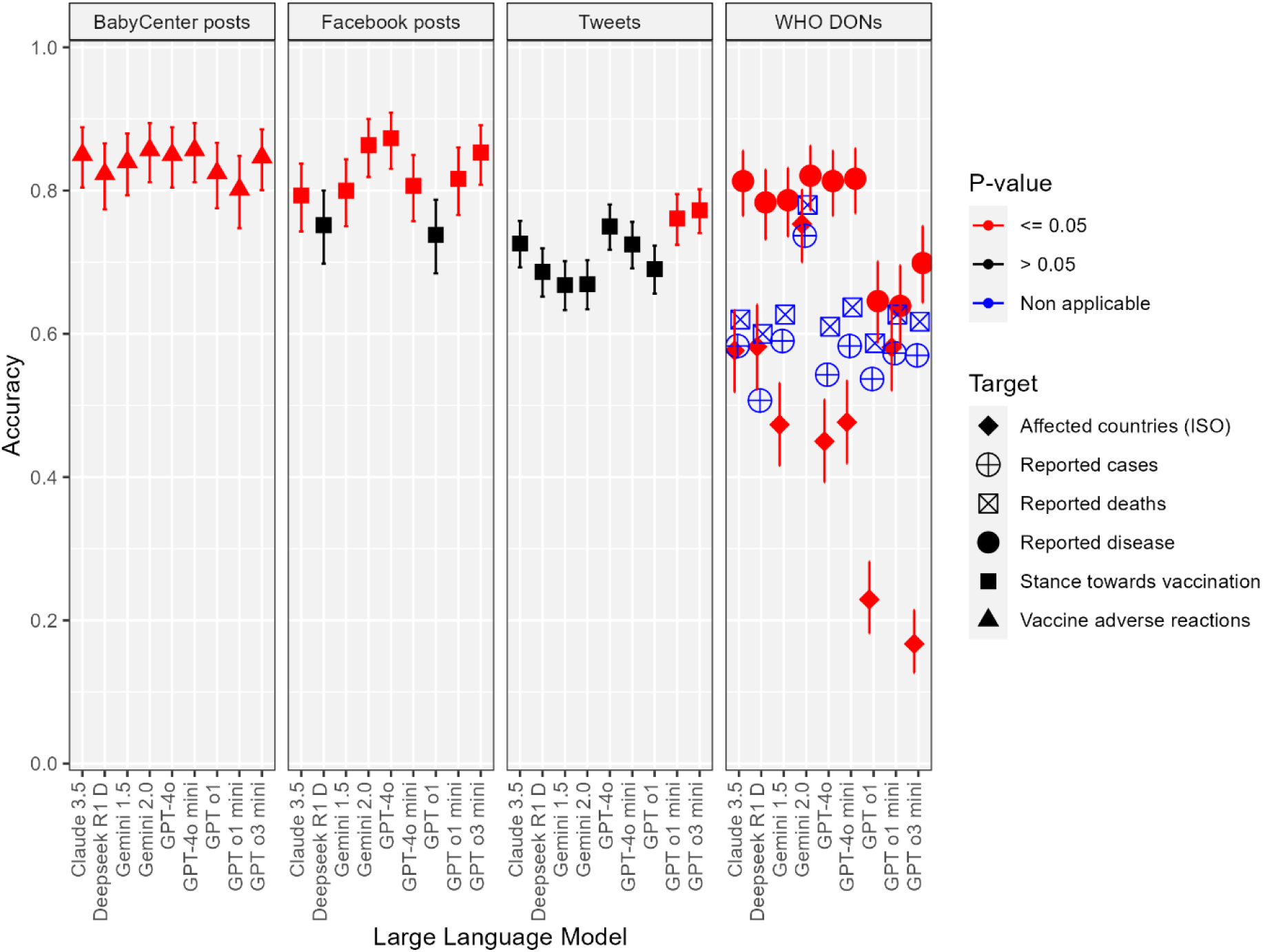
Accuracy for all target variables and 95% confidence interval for the numerical target variables per experiment, target and LLM.

In extracting epidemiological information from WHO DONs, after manually aligning the nomenclature, the reported diseases were extracted with similar accuracy for most LLMs, except for GPT o1, o1 mini and o3 mini which had lower values. The accuracy for extracting affected countries using three-letter ISO codes varied among LLMs. Numerical variables (reported cases and deaths) had a consistent accuracy across LLMs. Gemini 2.0 showed the highest accuracy for extracting affected countries, and reported cases and deaths.

#### 3.3.2. Classwise evaluation of categorical variables for social media and forum

Figure 4 shows the classwise evaluation of categorical variables for social media and forum use cases, including balanced accuracy, sensitivity, specificity and F1 score for each use case, target variable and class. Overall, the balanced accuracy, sensitivity, specificity and F1 score was different per each class in most of the cases, but the prevalence of each class in the predictions was very similar or equal to the prevalence of each class in the labelled dataset.

**Figure 4.**
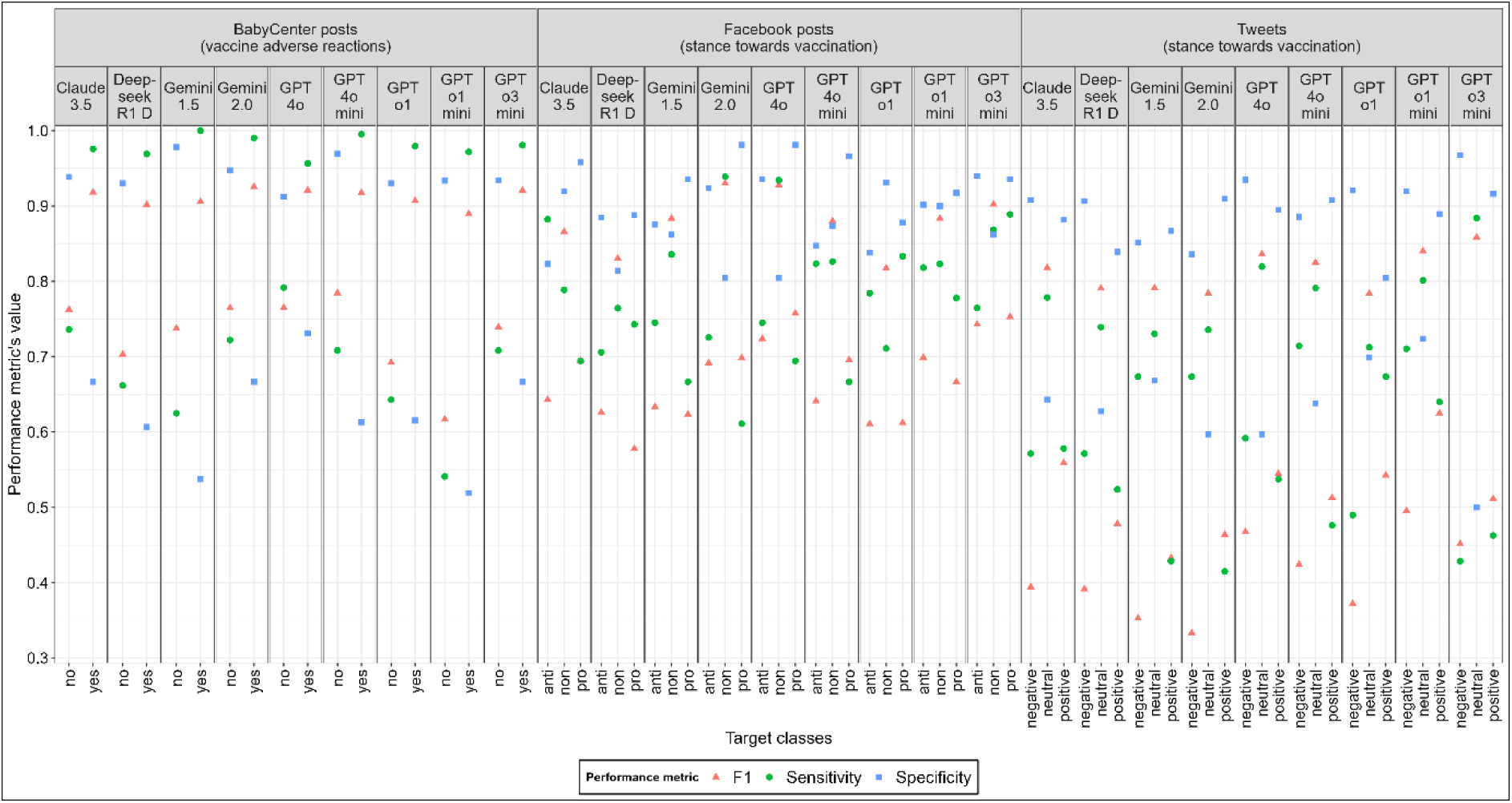
F1-score, sensitivity and specificity per experiment, LLM and class for BabyCenter post, Facebook posts and tweets.

In the predictions of vaccine adverse reactions in BabyCenter posts, the balanced accuracy was similar among classes. Sensitivity and F1 score were higher for the majority class (“yes”), while specificity was higher for the minority class (“no”).

For Facebook stance classification, classwise performance metrics were generally similar across classes, with higher F1 score for the majority class (“non”). For Twitter stance classification, the majority class (“neutral”) had consistent metrics across LLMs, showing the highest sensitivity and F1 score and lowest specificity. Classwise performance metrics for the other two classes (“positive” and “negative”) varied across LLMs.

#### 3.3.3. Classwise evaluation of categorical variables for WHO DONs

Due to the large number of unique classes for affected countries and diseases/pathogens, classwise performance metrics (balanced accuracy, sensitivity, specificity and F1 score) were calculated only for those classes within the third quartile of frequency in the dataset (Fig 5).

**Figure 5.**
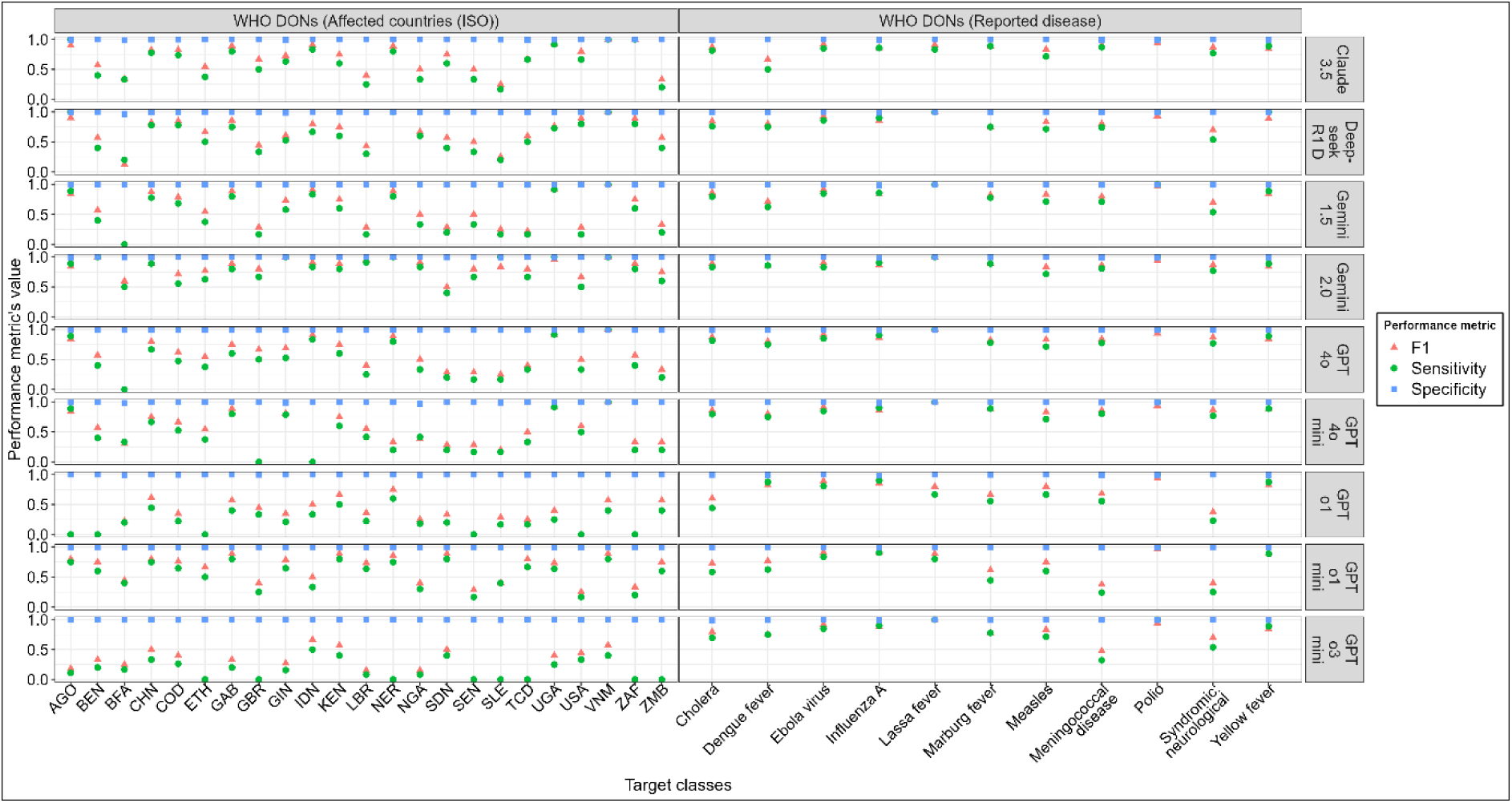
F1-score, sensitivity and specificity per experiment, LLM and class for WHO DONs.

Similarly to the other public health use cases, the prevalence of each class in the predictions was very similar or equal to the prevalence of each class in the labelled dataset.

In the predictions of diseases/pathogens, balanced accuracy was generally consistent across classes, except for Lassa fever and polio which had a 100% balanced accuracy, and GPT o1 and o1 mini which provided lower balanced accuracies across all classes. Sensitivity, specificity and F1 score were similar for most diseases/pathogens and GPT o1 and o1 mini produced lower values.

In the predictions of affected countries, balanced accuracy differed by country and LLM with a median of 75% (IQR 50% - 99.8%). GPT-4o mini, o1 and o3 mini showed the lowest balanced accuracy values. Countries with lower prevalence in the dataset had lower balance accuracy values.

Despite requesting LLMs to provide the affected country using the three-letter ISO (International Organization for Standardization) code, there were some predictions with the full name of the country, two-letter ISO code or other abbreviations used (e.g., DRC instead of COD for the Democratic Republic of the Congo).

## 4. Discussion

In this study, we have presented an open-source solution comprising two prototypes for LLM prediction, evaluation and benchmarking of public health use cases. We have shown their applicability and effectiveness in assessing LLM performance across categorical and numerical variables by testing these prototypes on four different public health use cases.

The open-source solution was designed to address the lack of public-health-specific standards for LLM benchmarking and the lack of an automated and near-real-time open-source solution to facilitate this task in the context of having recurrently new LLMs being released. The LLM prediction prototype requires standardised inputs and provides structured outputs ensuring consistency in the evaluations and includes different popular LLMs for broad comparison. The Shiny app prototype enables automated evaluation of LLM predictions providing an accessible and transparent process for public health professionals.

While existing frameworks such as HELM and Evidently-AI offer robust capabilities, including prompt templating, standardized JSON output, built-in evaluation metrics, and extensible task definitions; these tools are primarily designed for technical users and general-purpose tasks. In contrast, our system targets public health practitioners and researchers who may not have programming expertise, offering a lightweight, end-to-end solution for benchmarking LLMs in domain-specific use cases.

Our system supports structured outputs, customizable prompts, and flexible inputs, with a Shiny app interface that enables use without advanced technical skills. It allows direct upload of domain-specific datasets through a graphical interface, configuration and reuse of prompts stored alongside predictions, and harmonized JSON outputs across LLMs. The evaluation interface includes automated computation of performance metrics and visualizations for both categorical and numerical variables. The outputs are structured with consistent identifiers and metadata, allowing them to be easily used in expert review, manual annotation, or integration into downstream tools used in public health data pipelines. These features are designed specifically to reduce technical barriers, promote reproducibility, and align with the unique requirements of public health data, intending to be accessible to non-technical users and provide domain relevance that complement, rather than replicate, existing large-scale evaluation frameworks.

Our analysis has shown variability in the LLM predictions depending on the task, dataset and LLM. For the two use cases on stance towards vaccination from social media platforms, we observed a higher proportion of negative stance posts in Facebook and was not the minority class as in tweets. This discrepancy was likely due to the dataset used from both platforms, since half of the Facebook posts originated from pages considered to be contrary to vaccination. Moreover, the lower accuracy for tweets can be explained by the random selection of tweets, not originating from pages considered favourable or contrary to vaccination, which may be inherently less objective, even for human annotators as shown in a similar study [6].

For the WHO DONs, the accuracy was overall lower than for other use cases, except for reported disease after manual harmonisation of the terminology. Some of the WHO DONs had epidemiological features for more than one country which led to some LLMs to select the firstly reported or considered more relevant and, for other LLMs, to include the epidemiological information also for the countries that were not the main focus of the WHO DON. These underlies, on the one hand, the need for integrating public health ontologies and structured terms (e.g., International Classification of Diseases -ICD-) to enable semantic interoperability for the reported diseases and a clearer and more explicit prompt specifying the most relevant information which is expected to be extracted. Despite providing such structured terminology for the affected countries, there were still inconsistencies which could be handled by explicitly pointing out to a reference with the list of countries and corresponding two-letter ISO code. These discrepancies could have also taken place by human annotators if such references and explicit instructions were not provided either.

In relation to the last public health use case in detecting vaccine adverse reactions from BabyCenter forum posts, the accuracy was higher compared to the stance towards vaccination. This can be explained by the inherent objectivity in detecting this information and by having two clearly defined classes (i.e., yes and no), whereas stance classification involves a degree of interpretation and agreement has not been fully reached even with human annotators [6]. Despite this may suggest that LLMs may have higher performance in tasks that require objective and structured information extraction, tasks requiring subjective interpretation or classification can benefit also from the use of LLM if prompt engineering and understanding of the dataset is performed.

Our findings showed that no LLM consistently outperformed others across all tasks and use cases, emphasizing the need for selecting task- and use-case specific models. Even for the same task (e.g., stance towards vaccination from social media posts), having different dataset composition impacted the performance of the same LLMs. Furthermore, smaller models outperformed larger counterparts in various cases. For example, this was the case for GPT o1 mini in predicting stance towards vaccination from Facebook posts or GPT-4o mini in predicting affected countries and reported cases or deaths in WHO DONs. This suggests that the complexity of the LLM alone does not determine effectiveness and that other aspects should also be considered, such as computational efficiency and costs, particularly for resource-constrained settings.

Furthermore, our results reinforce that non-domain-specific LLM benchmarks may not be directly applicable to the use of LLMs in public health use cases. While the models included in this study are expected to have a high performance overall, their performance was not as high in domain-specific tasks or specific use cases such as the extraction of epidemiological data. Likewise, when comparing the performance of LLMs from the same company (e.g., OpenAI models), the latest models show higher performance for the general tasks for which these are being evaluated [20]. However, we have seen that, for example, GPT o3 mini does not always have the highest performance values when compared with the other GPTs included in this study.

While this study presented a structured approach for LLMs prediction, evaluation and benchmarking in public health, it has some limitations. The LLM prediction prototype requires deployment which may need the involvement of additional technical expertise; however, to mitigate this, the Shiny app allows other input files with LLM predictions obtained using other solutions. Furthermore, while the Shiny app was only tested in Windows, R Shiny is expected to function similarly across other operating systems. Although not all available LLMs were included in our benchmark, we ensured to have a representative mix of popular models with reported high performance. Further LLMs and public health use cases can be added in the future to further assess the usability of this tool. We have not conducted a comprehensive usability assessment of the open-source solution; however, its development was guided by simplicity, user-friendliness and accessibility principles from the perspective of public health professionals.

Furthermore, both prototypes are accompanied by public repositories containing detailed usage instructions and fully documented code. These features are intended to support ease of use, though we acknowledge that formal usability evaluation (e.g., surveys or user testing) would be a valuable step in future work to assess and validate the system’s accessibility for public health practitioners.

This study contributes to the effective integration and use of LLMs in public health by providing a structured, user-friendly and scalable solution for LLM prediction, evaluation and benchmarking. Our results underline the relevance of standardised and task-specific evaluation methods and model selection, and the use of clear and structured prompts to improve LLM performance in domain-specific use cases. Future studies should evaluate its real-world usability and adoption in public health, and considering the fast-paced evolution of LLMs and other use cases requiring images, expand the solution incorporating additional models to ensure up-to-date assessments and testing its usability for LLM predictions based on images.

## Authors contributions

LE was responsible for conceptualization, data curation, formal analysis, methodology, development of the Shiny app prototype for LLM evaluation and benchmarking, visualization, and writing original draft manuscript. MS was responsible for conceptualization, funding acquisition, methodology and supervision. DK was responsible for developing the LLM prediction prototype. SC was responsible for the public health use case on extracting relevant features from WHO DONs. KK and YM were responsible for the public health use cases on stance towards vaccination in Facebook posts and detecting vaccine adverse reactions from BabyCenter US forum. All authors contributed to the editing of the final manuscript, and have seen and approved the manuscript.

## Supporting information

**S1 Text. Prompts used for the public health use cases.**

