## Supporting information (S1 Text) for "Open-source solution for evaluation and benchmarking of large language models for public health"

### S1 Text. Prompts used for the public health use cases.

#### 1. Stance towards vaccination in English tweets

Give me the stance of the user towards vaccination in the tweet text, regardless of the general sentiment expressed in the tweet. A tweet with a negative general sentiment can express a positive stance of the user towards vaccination and viceversa. \ Use one of the following words: neutral, negative, positive. Include an explanation for the selection of the stance. \ The reply must be a json file with the following keys: stance (only 'neutral', 'positive' or 'negative'), explanation. \\ News, factual or objective tweets as well as generally ambiguous examples should be classified as neutral except if:\ * User agrees with a positive statement on vaccines or vaccinations which should be classified as positive \ * User disagrees with a positive statement on vaccines or vaccinations which should be classified as negative \ * User is angry or not happy when vaccines are not available which should be classified as positive \ * User is angry or not happy with a negative statement on vaccines or vaccinations which should be classified as positive \ * User is happy with a negative statement on vaccines or vaccinations which should be classified as negative \ * User is enquiring for availability or increased availability of a vaccine which should be classified as positive \\ Classify as positive if:\ * The user expresses hope or anticipation towards a vaccine.\ * The user expresses desire for having more vaccine doses. \ * The user is ironic on people being negative towards vaccine (e.g., I am not getting a vaccine because they are microchipping you. Posted on social media , on a phone..that has all your info, your location...and the info is being shared and sold ...Ummmm ok) \\ Examples of positive tweets: \ * Some news of hope. Hopefully this vaccine becomes success and India becomes leader again. \ * This has been a horrendous year for so many people who have lost loved ones. It would be wonderful if the #vaccine was effective and could be administered world-wide to prevent further loss. My fingers and toes are crossed. \ * A healthy recovery of the world from COVID-19 is only possible when no one is left behind making it necessary that the vaccine is accessible to all without any hurdle. #HealthForAll #Equality #Equity #publichealth #awareness.\ * IF I SEE ONE MORE FUCKING POLITICIAN GET THE VACCINE BEFORE FRONT LINE WORKERS I'M GONNA LOSE MY SHIT!! \ * When will we learn to respect our scientific community ? Pathetic that ppl want to play politics on the vaccine \ * @user @user @user etc if U have so much problem so plz don't get them vaccinated, we know, U have enough money for treatment. U guys fall mild sick, immediately rush to medanta hospital.\ * $MRNA received approval across the pond in the U.K. demand increasing globally for a vaccine as cases rise. #stocks #finance #markets #investing #money #wealth #economy #OptionsTrading \ * On the other hand, they're more likely to receive much more effective and safe vaccines, so it's not quite as bad as you might think \\ Examples of neutral tweets: \ * AstraZeneca Pauses Vaccine Trial After Test Subject Gets Ill <url> \ * The federal government outlined a sweeping plan Wednesday to make vaccines for COVID-19 available for free to all Americans. <url> \\ Examples of negative tweets: \ * There's a reason these vaccines can take up to 20 years to reach market. Say no to the vaccine. \ * Vaccines may not be good for kids like the Dengue Fever vaccine. You’re playing with fire. <url> \ \

### 2. Stance towards vaccination in Facebook comments

Give me the stance of the user towards vaccination in the Facebook text, regardless of the general sentiment expressed in the text. A text with a negative general sentiment can express a positive stance of the user towards vaccination and viceversa. \ Use one of the following words: NON (for neutral stance), ANTI (for negative stance), PRO (for positive stance). Do not include any explanation and do not invent. \ The reply must be a json file with the following key: stance (only 'NON', 'ANTI' or 'PRO'). \ \

### 3. Detecting vaccine adverse reactions from BabyCenter US forum

Please indicate whether the text extracted from a forum entitled 'baby center' includes any adverse reaction or side effect to vaccination and, if yes, which adverse reaction or side effect. \ Do not invent. \ Format your response as a JSON object with the following keys: class (yes/no), adverse reaction (free text, preferable very few words referring only to the adverse reaction or side effect).

### 4. Extracting relevant features from WHO DONs

From the text extract the following items:\ 1 - The name of the virus that has caused the outbreak.\ 2 - The three-letter ISO code of the country where this virus outbreak occurred, if present. Do not invent. \ 3 - The date when this virus outbreak occurred, if present. Do not invent.\ Show the date in the format YYYY-mm-dd.\ 4 - The number of cases derived exclusively from the virus outbreak mentioned in the text, if present. Do not invent \ 5 - The number of deaths caused exclusively by the virus outbreak mentioned in the text, if present. Do not invent.\ Format your response as a JSON object with the following keys: virus, country, date, cases, deaths. \ If the information is not present, do not invent and use "NA" as the value.\ Write no explanations or notes . \
